# Candidate gene modifiers of dystrophinopathy identified by the uniform application of genome-wide datasets to novel GWAS-identified loci

**DOI:** 10.1101/2021.11.03.21265899

**Authors:** Kevin M. Flanigan, Megan A. Waldrop, Paul T. Martin, Roxane Alles, Diane M. Dunn, Lindsay N. Alfano, Tabatha R. Simmons, Melissa Moore-Clingenpeel, John Burian, Sang-Cheol Seok, Veronica J. Vieland, Robert B. Weiss

**Author notes:** Corresponding Author: Kevin M. Flanigan, MD, Center for Gene Therapy, Nationwide Children’s Hospital, 700 Children’s Drive, Columbus, OH 43209.

## Abstract

Although the major determinant of disease severity in patients with severe Duchenne muscular dystrophy (DMD) or milder Becker muscular dystrophy (BMD) is whether their dystrophin gene (*DMD*) mutation disrupts the mRNA reading frame or allows expression of a partially functional protein, other genes have been proposed or demonstrated to modify the severity of disease progression. In a companion paper to this one, we describe our novel approaches to genome-wide association study (GWAS) of loss of ambulation (LOA) in the largest genome-wide search to date for loci influencing disease severity in DMD patients. Candidate regulatory SNPs that modify disease progression were identified using an evidential statistical paradigm and here we present a uniform application of recent functional genomic datasets to explore the potential functional impact of the top six candidate regions with PPLD scores of ≥0.4. The results of this analysis of the largest DMD GWAS survey to date elucidate recurrent and potentially new pathways for intervention in the dystrophinopathies.

## Introduction

Mutations in *DMD*, encoding the dystrophin protein, result in the X-linked dystrophinopathies. The most common of these are the severe Duchenne muscular dystrophy (DMD) and the milder Becker muscular dystrophy (BMD), with some patients demonstrating an intermediate phenotype. The primary determinant of disease severity depends upon whether the mutation truncates the *DMD* open reading frame, ablating dystrophin expression, or maintains an open reading frame, allowing expression of a partially functional muscle-specific (Dp427m) dystrophin isoform.

Although treatment with corticosteroids or environmental factors may alter the age of loss of ambulation (LOA) in patients who lack dystrophin expression, increasing evidence implicates other genetic modifiers of severity, including proteins involved in inflammation, fibrosis, regeneration, and sarcolemma repair ^1-3^. Protein polymorphisms in latent TGF-β binding protein 4 (*Ltbp4*) are associated with increased levels of TGFβ-mediated fibrosis in mice^4^, and a common human *LTBP4* haplotype has been associated with disease severity. The matricellular protein osteopontin (*Spp1*) contributes to the balance between inflammatory, regenerative, and fibrotic processes in muscle, and common regulatory SNPs in the proximal promoter of *SPP1* are associated with variations in age at LOA in DMD patients ^5^. Cohort effects of *SPP1* and *LTBP4* SNPs are evident with variable success at replication ^2^, but the effects of the “protective” and “risk” human LTBP4 polymorphisms on the dystrophic phenotype in mice have been confirmed, with epistatic interactions between *Spp1* and *Ltbp4* in modulating fibrosis via pathological TGFβ signaling ^6^. The sarcolemmal repair protein and downstream effector of TGFβ signaling annexin A6 (*Anxa6*) modulates severity in mice, with alleles of *Ltbp4* and *Anxa6* acting jointly to modify the dystrophic phenotype ^6^. The Notch signaling pathway gene *Jagged1* modifies severity in the golden retriever muscular dystrophy (GRMD) dog model, implicating a role for satellite cell self-renewal and quiescence in severity^7^.

Prior GWAS studies used different phenotypic classification schemes and statistical analyses than what we describe here. The first, utilizing an exome chip in 109 ancestrally homogeneous sample implicated a functional SNP (rs1883832) in the Kozak sequence of the *CD40* gene ^1^, which encodes a costimulatory receptor mediating immune and inflammatory responses. The second, from our group, was a preliminary GWAS using 253 non-ambulant subjects ^3^ in which we observed two loci above the genome-wide significance threshold using the recessive model. One locus was *LTBP4* itself, where fine mapping revealed *cis*-eQTL variants in linkage disequilibrium with “protective” IAAM LTBP4 protein isoform and associated with *LTBP4* mRNA expression. The other locus was a distal enhancer containing *cis*-eQTL variants associated with expression of thrombospondin-1 (*THBS1*), encoding a matricellular protein with multifunctional properties including promoting TGFβ signaling through latent protein complex activation. Finally, a recent two-stage design using whole exome sequencing of extreme phenotypes identified *TCTEX1D1* as an LOA modifier ^8^. Such studies highlight the importance of novel study designs in searching for modifiers of DMD.

As described in a companion paper [citation], we have performed a GWAS analysis on N=419 dystrophinopathy patients using a different approach. Specifically, we used a *novel and conservative approach to classifying DMD genotypes* to minimize the potential influence of residual dystrophin expression on phenotype, used *a new form of survival analysis* residual to evaluate how unexpected is an individual’s age at loss of ambulation, and used *an evidential statistical paradigm* to identify candidate SNPs that modify disease progression. Here we describe the *uniform application of recent genome-wide datasets* to that candidate SNP data to reveal their functional impact. Beginning with six candidate loci identified in the GWAS study, we were able to implicate several novel regulatory loci as potential modifiers of DMD severity. Utilizing a systematic *in silico* pipeline integrating ENCODE genomic annotations, chromatin interaction experiments, expression quantitative trait loci (eQTL), and *in silico* functional predictions, we consider the biological plausibility of these candidate modifiers as a first step toward further studies to determine pathogenesis.

## Materials and Methods

The study design is described in detail in the companion paper, as is the subject cohort [Flanigan et al., medXriv, 2021]. GWAS was based on the posterior probability of (trait-marker) linkage disequilibrium (PPLD) to assess evidence for or against association between LOA and each SNP in turn. The rationale for the selection of and the advantages of the PPLD over regression analysis in the context of GWAS based on small to moderate sample sizes are detailed elsewhere ^9^. The methods of (i) functional annotation and eQTL analysis of candidate loci and (ii) analysis of genomic regulatory features are included in the Supplemental Methods.

## Results

A PPLD threshold ≥ 0.05 defined credible sets from the top 6 regions, identifying 96 total SNPs for detailed analysis. Functional annotations of individual SNPs (Table S1) revealed that all 96 SNPs were noncoding intergenic or intronic variants. Table 1 shows the lead candidate SNP from each of the regions associated assigned to a candidate protein coding gene using the Open Target Genetics V2G pipeline with the gene expression data (eQTL) supporting this connection. These six credible SNP sets with a PPLD score ≥ 0.4 (Table 1) were non-coding and therefore we examined these genomic regions for biological features indicative of functional regulatory variation. These included ENCODE candidate *cis*-regulatory elements (cCREs), ENCODE indexed DNase I hypersensitive sites (DHSs), ENCODE transcription factor footprints, gene proximity, enhancer-promoter chromatin interactions, skeletal muscle total RNA expression, and *cis*-eQTLs SNP-gene pairs from the GTEx and the eQTL Catalogue databases.

**Table 1.** Top six regions with PPLDs ≥ 0.40. rsID is the top genotyped SNP per region, where the regions are defined as in the main text. PPLD: The PPLD at the genotyped SNP. MAF: minor allele frequency in study subjects. Overall V2G top gene: Variant to Gene nearest protein coding gene. eQTL: gene expression data (eQTL) supporting this connection.

| Chr | rsID | Gencode Category | PPLD | MAF | Overall V2G (distance to canonical TSS) | eQTL SNP-to-gene | Tissue | eQTL p-value |
| --- | --- | --- | --- | --- | --- | --- | --- | --- |
| 2 | rs34263553 | intergenic | 0.87 | 0.13 | ETAA1 (334,439 bp) | ETAA1 | T cells | 1.70E-05 |
| 4 | rs1358596 | ncRNA_intronic | 0.75 | 0.19 | GALNTL6 (301,109 bp) | GALNTL6 | T cells | 7.20E-04 |
| 5 | rs12657665 | intergenic | 0.62 | 0.48 | ADAMTS19 (76,055 bp) | ADAMTS19 | Sk. Muscle | 9.70E-04 |
| 6 | rs10499096 | intergenic | 0.64 | 0.05 | MAN1A1 (820,646 bp) | - | - | - |
| 8 | rs3899035 | intergenic | 0.83 | 0.11 | NCALD (53,501 bp) | NCALD | Monocytes | 4.30E-14 |
| 18 | rs2061566 | upstream;downstream | 0.40 | 0.09 | PQLC1 (159,919 bp) | PQLC1 | Neutrophils | 2.70E-16 |

### Chr2 rs34263553 to ETAA1 connection

#### Credible SNP set, eQTLs, and chromatin features

The nearest protein-coding gene to the chr2 rs34263553 SNP with the highest PPLD value (0.87) is the Ewing tumor-associated antigen 1 (*ETAA1*) gene, at a distance of 335 kb (Figure 1A). One-to-all Hi-C chromatin and pCHi-C data support a physical interaction between the rs34263553 region and the *ETAA1* promoter region in psoas muscle (Figure 1A and Table S3). This region also coincides with a *cis*-eQTL data associated with *ETAA1* expression in T cells (Figure 1A and Table S2). The credible rs34263553 SNP interval contains multiple cCREs with both distal enhancer-like and CTCF-only signatures (Figure 1B) and the lead SNP rs34263553 is also located within a stromal B index DHS (Table S4), flanked upstream by an enhancer cluster and downstream by a CTCF-binding site with a prominent DHS peak in multiple tissues/cells (Figure 1B). The 12 SNPs in the rs34263553 PPLD credible set coincided completely with the 13 SNP fine mapped *ETAA1* eQTL credible set defined by the SuSiE method and rs34263553 had the highest posterior probability of being the causal eQTL variant for *ETAA1* expression (Figure 1C). Regulatory features in the vicinity of rs34263553 include the phylogenetically conserved enhancers, the strong CCCCTC-Binding factor (CTCF) site active in many cell types, and a nearby binding site for FOXA1/A2 pioneer transcription factor (Figure 1C). In the mouse, *Etaa1*-deficient T cells display elevated DNA damage during proliferation ^10^ and the negative beta value in T cells for the *cis*-eQTL suggests that the minor allele of rs34263553 is associated with decreased *ETAA1* expression in humans.

**Figure 1.**
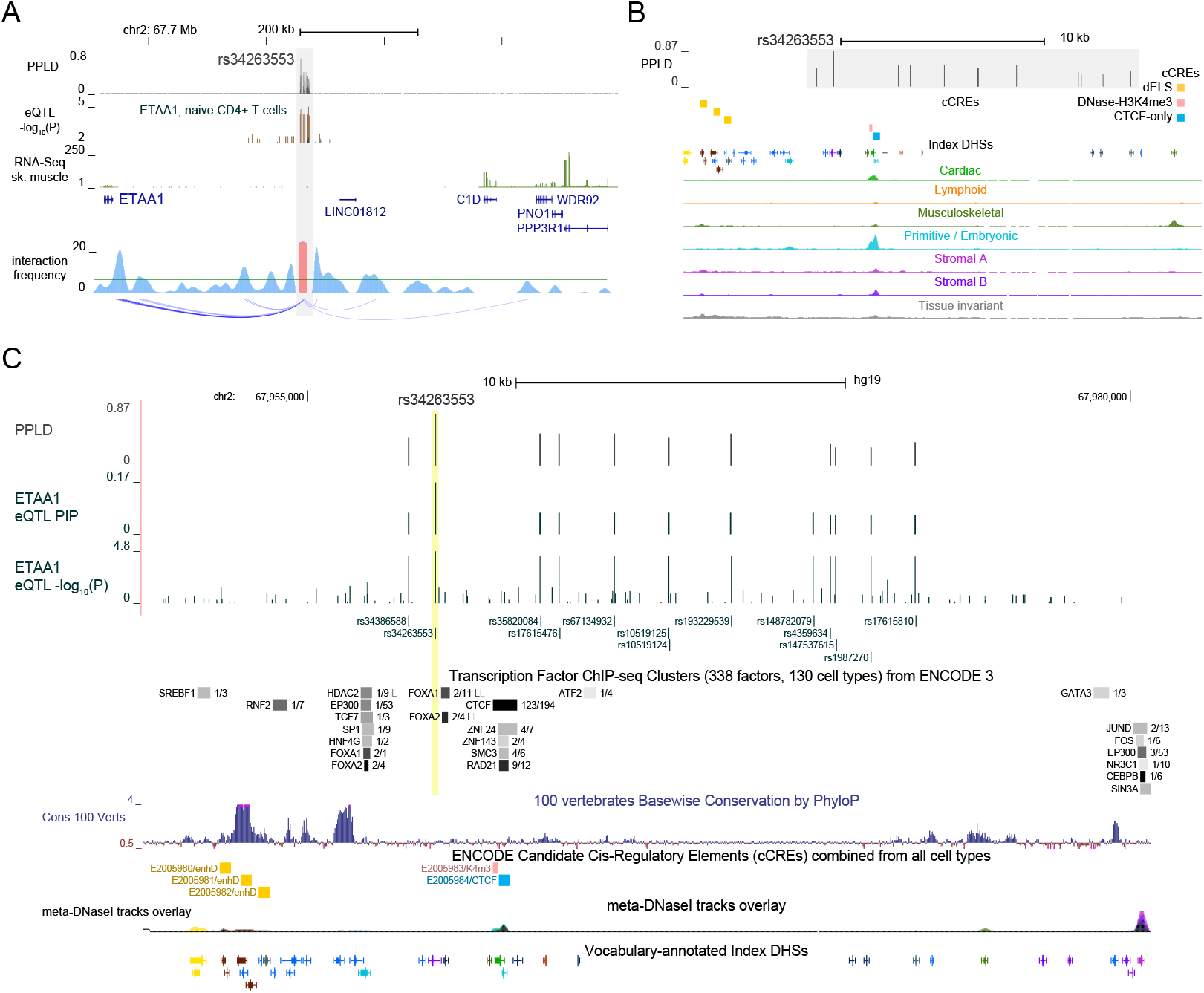
eQTL and chromatin features for the chr2 SNP region. (A) chr2:67,600,001-68,560,000 (hg19) including PPLD values for genotyped and imputed SNPs, total RNA-Seq read depths from pooled normal skeletal muscle tissue, RefSeq Select transcripts, and SNP-*ETAA1* eQTL p-values from T cells (Table S2). The grey bar highlights the PPLD credible SNP set region and the lead SNP rs34263553. The bottom track is a one-to-all Hi-C chromatin interaction plot from psoas muscle where the y-axis (blue density) indicates bias-removed interaction frequencies, and the horizontal green line is a cut-off value (=2) for the distance-normalized frequency. (B) The credible SNP region (grey bar) from (A) with annotation tracks for ENCODE cCREs, vocabulary-annotated index DHSs elements, and biologically labeled component meta-DNase I tracks from the top 15 ENCODE biosamples most strongly associated with each component. (C) Overlap of the chr2 PPLD credible SNPs and lead SNP rs34263553 with the *ETAA1*-eQTL from (A) shown as both the nominal -log_10_ p-value of the eQTL association and the eQTL fine mapped using SuSiE to assign the posterior inclusion probability (PIP) to the SNPs in the eQTL credible set. Additional detail is shown for the ENCODE cCREs features, as well as conserved sequence elements from vertebrate alignments scored with PhyloP.

#### ETAA1 as a plausible modifier gene

ETAA1 is an activating protein for ATR, the major kinase regulating replication stress and the DNA damage response pathways ^11^. The ETAA1-ATR complex has a role in monitoring the completion of S phase and in regulating the S/G2 cell cycle transition^12^. *Etaa1*-deficient mice ^10^ have defects in processes requiring rapid cell proliferation, including deficiencies in clonal T cell expansion and incompletely penetrant embryonic lethality. Rapid proliferation is a requirement for muscle stem cells (MuSCs) to participate in repair and regeneration of damaged myofibers caused by the loss of dystrophin. ETAA1-dependent activation of ATR during mitosis has also been shown to be important for Aurora B kinase (AURKB) signaling ^13^ and in preventing chromosomal misalignments during metaphase. A plausible role for ETAA1 as a modifier would be in maintaining genome integrity and chromosomal stability of the muscle stem cell population.

### Chr4 rs1358596 to GALNTL6 and chr6 rs10499096 to MAN1A1 connections

#### Credible SNP sets, eQTLs, and chromatin features

*GALNTL6* is the nearest protein-coding gene to the chr4 rs1358596-tagged region and encodes one of twenty known polypeptide O-GalNAc transferase genes. The credible SNP region overlapped a known *GALNTL6* eQTL interval and several HiC interactions connect this region to the *GALNTL6* promoter. One-to-all Hi-C chromatin (Figure 2A) and promoter-capture Hi-C interactions link the chr4 SNP region to the *GALNTL6* promoter region and the lead SNP rs1358596 also coincides with a SNP block observed as a *cis*-eQTL for *GALNTL6* expression in T cells in a region overlapping divergent ncRNA promoters (Fig 2A). The rs10499096-tagged chr6 SNP region is linked through long range promoter-capture Hi-C (pCHi-C) interactions in muscle tissue to the nearest protein-coding gene *MAN1A1*, which along with *MAN1A2* and *MAN1C1*, encode a family of Golgi α1-2-mannosidases. One-to-all and pCHi-C interactions (Figure 2C and Table S3) link the chr6 SNP region to the neighboring promoter region of the *MAN1A1* gene. The lead SNP rs10499096 is located ∼820 kb upstream of the *MAN1A1*and long-range pCHi-C interactions are observed between the SNP region and the *MAN1A1* promoter in 16 different tissues/cell types (Table S3). The credible SNP region includes a prominent musculoskeletal DNase hypersensitive site coincident with cCRE CTCF-binding sites (Figure 2D), suggesting a functional link to altered chromatin looping.

**Figure 2.**
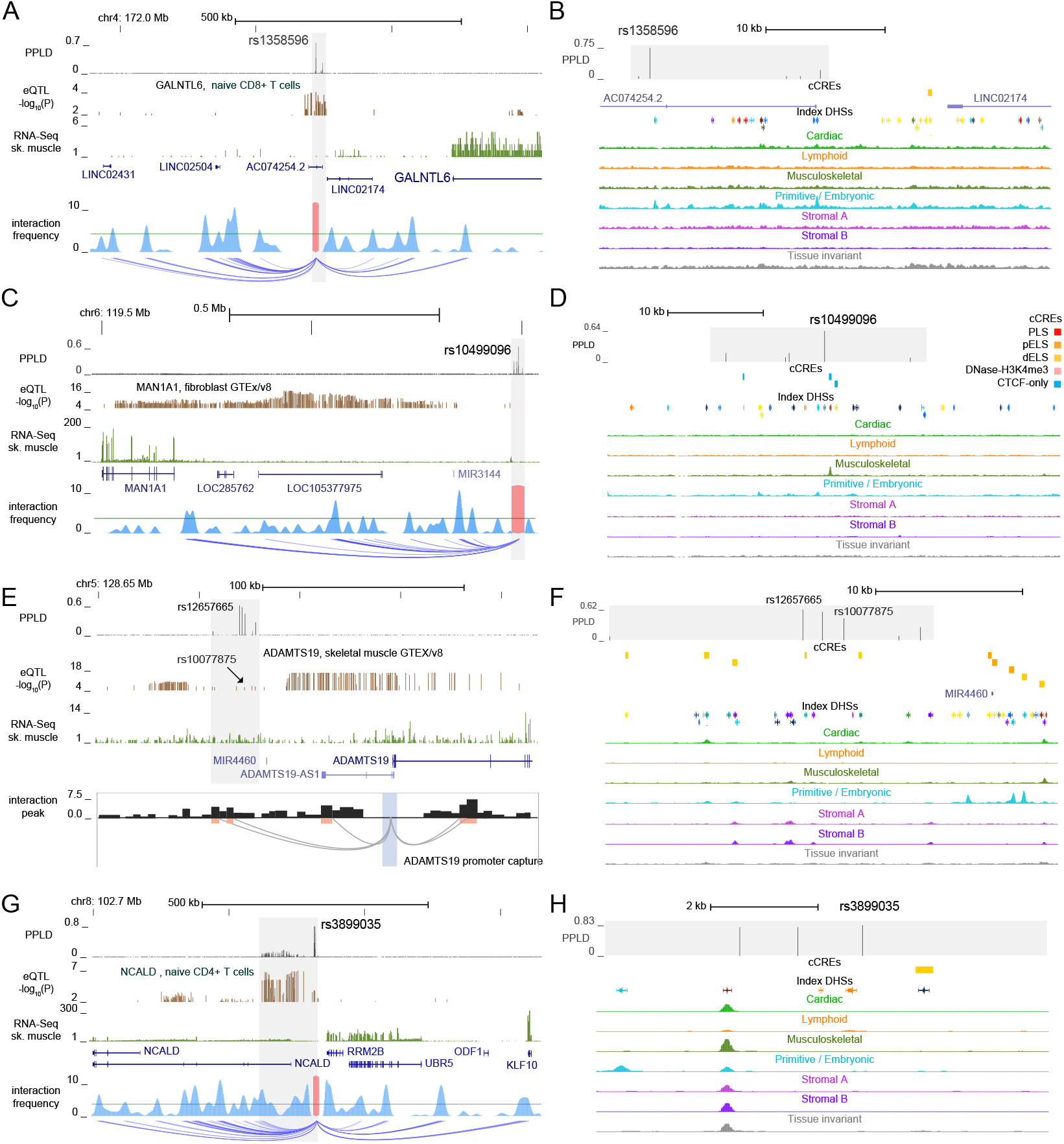
eQTL and chromatin features for the chr4, chr6, chr5, and chr8 SNP regions. (A) The lead SNP rs1358596 from chr4:171,920,001-172,940,000 (hg19) with annotation tracks for PPLD values, SNP-*GALTNL6* eQTL p-values from T cells (eQTL catalogue, Table S2), skeletal muscle total RNA-Seq read depths, RefSeq Select transcripts, and the PPLD credible SNP region highlighted by the grey bar. The bottom track is a one-to-all Hi-C chromatin interaction plot from psoas muscle. (B) The credible SNP region (grey bar) from (A) with annotation tracks for ENCODE cCREs, vocabulary-annotated index DHSs elements, and biologically labeled component meta-DNase I tracks as in Fig. 1. (C,D) Similar annotation tracks as in (A,B) from chr6:119,480,001-120,540,000 with the lead SNP rs10499096 including PPLD values for genotyped and imputed SNPs, SNP-*MAN1A1* eQTL p-values from GTEx v6 fibroblasts, skeletal muscle total RNA-Seq read depths, RefSeq Select transcripts, and highlighted credible SNP region. The bottom track is a one-to-all Hi-C chromatin interaction plot from psoas muscle. (E,F) Similar annotation tracks as in (A,B) from chr5:128,640,001-128,870,001 with the lead SNP rs12657665 and the SNP-*ADAMTS19* eQTL p-values from GTEx v8 skeletal muscle. The bottom track in the left panel shows promoter capture Hi-C chromatin interactions from cardiomyocytes using the *ADAMTS19* chr5:128,789,920-128,798,524 promoter fragment (see Table S3). Barplots are normalized promoter-captured chromatin interactions (y-axis = -log10 p-values) linked to the *ADAMTS19* promoter (light blue). Nominally significant chromatin interactions are shown as grey loops linked to their distal HindIII fragments highlighted in orange. (E,F) Similar annotation tracks as in (A,B) from chr8:102,685,001-103,690,000 with the lead SNP rs3899035 and the SNP-*NCALD* eQTL p-values from BLUEPRINT T cells.

#### Glycosylation genes as plausible modifiers

The *GALNTL6* gene produces a protein that glycosylates the EA2 peptide once it has already been modified with O-GalNAc at another site by *GALNAC-T1*^14^. Thus, *GALNTL6* may be capable of increasing O-GalNAc glycan density on proteins but not initiating glycosylation in brain, spinal cord, trachea, testes and skeletal muscle^14^. A GWAS study comparing endurance athletes and sedentary controls identified marker rs558129 in the *GALNTL6* locus by meta-analysis (p=0.0002) ^15^. Interestingly, while elite endurance athletes were associated with the rs558129 (C), a study comparing 49 Russian strength (weight-lifters and power-lifters) athletes, 169 endurance athletes, and 201 controls found that the rs558129 (T) was more concentrated in the strength athletes relative to endurance athletes (p=0.0067) or controls (p=0.0036)^16^. Thus, *GALNTL6* may dictate a switch for muscles to be predisposed to endurance versus strength training.

Altered *MAN1A1* expression could theoretically impact muscle ion channel function, muscle cell adhesion, and muscle stem cell function. For example, removal of sialic acids via lowered *MAN1A1* expression will increase high mannose N-linked glycosylation and could cause dramatic depolarizing shifts in NaV channels, as can occur when muscle cells are treated with neuraminidase, which removes sialic acids ^17^. Removal of muscle N-linked sialic acid can also remove hyperpolarizing shifts in the β1 chain of the NaV channel and can increase conductance of muscle voltage-gated calcium channels, which could impact calcium homeostasis, which is known to be altered in muscular dystrophy^18^. *KLOTHO* has been shown to ameliorate muscular dystrophy ^19^ and alter muscle regeneration and muscle mitochondria and stem cell function^20,21^. KLOTHO protein contains an α2,6 sialidase activity that also may reduce cell surface sialic acids ^22^. Thus, by altering N-linked sialic acid, changed *MAN1A1* expression could also mimic biological effects of *KLOTHO* in muscle.

### Chr5 rs12657665 to ADAMTS19 and chr8 rs3899035 to NCALD connections

#### Credible SNP sets, eQTLs, and chromatin features

The nearest protein-coding gene to the chr5 rs12657665-tagged region is *ADAMTS19* which encodes an ADAMTS (<u>a d</u>isintegrin-like <u>a</u>nd <u>m</u>etalloproteinase domain with thrombo<u>s</u>pondin-type 1 motifs) protease involved in extracellular matrix formation, interactions, and remodeling. The credible SNP region is embedded within an extended *ADAMTS19* skeletal muscle eQTL from the GTEx v8 database (Figure 2E), although the strongest eQTL signal is in weak LD with rs12657665 and rs10077875, suggesting a more specific dysregulation in dystrophic tissue. The credible SNP region contains multiple cCRE enhancers and prominent stromal/musculoskeletal DNase I hypersensitive site (DHS) peaks (Figure 2F), and contains multiple pCHi-C and *cis*-eQTL interactions with other neighboring genes (Tables S2 and S3). Notable cis-eQTLs associated with the lead SNP rs12657665 include *FBN2* encoding fibrillin-2, a component of extracellular microfibrils, and *CHSY3*, encoding a glycosyltransferase chondroitin sulfate synthase 3 located directly downstream from *ADAMTS19*.

The nearest protein-coding gene to the chr8 rs3899035 region is neurocalcin delta (*NCALD*), a myristoylated calcium sensor protein that shows calcium-induced binding to lipid bilayers. The credible SNP interval spans ∼120 kb, including a *NCALD* promoter with multiple SNPs intersecting distal enhancer-like cCREs regulatory elements. This region of chr8 has been previously identified as harboring regulatory variants that modify spinal muscular atrophy (SMA) disease progression via reduced *NCALD* expression improving endocytosis defects^23^. There are 67 SNPs in moderate LD with the lead intergenic rs3899035 SNP which flanks a prominent musculoskeletal DHS peak (Figure 2G and H). The moderate LD SNPs are annotated as a ‘super-enhancer’ (Table S1) and intersect 9 distinct cCRE enhancers, including rs17416694 located in the EH38E2653349 cCRE which has a prominent musculoskeletal DHS (Table S4). These SNPs overlap a *cis*-eQTL modulating *NCALD* expression in T cells, but other genes show *cis*-eQTLs in this region (Table S3) and are also supported by enhancer-promoter chromatin interactions (Figure 2G). These include *KLF10*, a zinc-finger Krüppel-like transcription factor activated as part of the early response to TGF-β signaling and whose absence in the *mdx* model exacerbates fibrosis ^24^.

#### ECM genes and NCALD as plausible modifiers

The ADAMTS19 protease belongs to a deeply diverged 19-member family with diverse functions associated with tissue morphogenesis, musculoskeletal development, and pathophysiological remodeling, and like LTBP4 and THBS1, is found in elastic fiber molecules, specifically fibrillin microfibrils ^25^. Recessive loss of function (LoF) mutations in *ADAMTS19* cause non-syndromic heart valve disease and the aortic valves from *Adamts19* knockout mice show a disorganized ECM due to defects in postnatal heart valve remodeling ^26^. Recessive LoF mutations that cause a rare connective tissues disorder known as Weill-Marchesani syndrome (WMS) are phenocopies of dominant pathogenic variants in fibrillin-1, *FBN1* (WMS2, OMIM 608328) and occur in *ADAMTS10* (WMS1, OMIM 277600), *LTBP2* (WMS3, OMIM 614819), and *ADAMTS17* (WMS4, OMIM 613195), a sister protease of ADAMTS19 ^27^. These acromelic dysplasias share phenotypes including pseudomuscular build, short stature, joint stiffness, brachydactyly, and tight skin. Recessive LoF mutations of *LTBP4* also cause a fibrillin-related connective tissue disorder, cutis laxa type 1C (OMIM 613177). It has been proposed that the biological substrate underlying these phenotypes is a protein network on FBN1 microfibrils that brings LTBPs and ADAMTS proteases into physical proximity to form a TGF-β signaling hub in the ECM ^28^. Although our current results did not identify previously seen associations with *LTBP4* and *THBS1*, this putative *ADAMTS19* association reinforces the role of TGF-β signaling dysregulation in modifying disease progression.

The Ca^2+^-sensor NCALD has been shown to regulate cyclic GMP synthesis from atrial natriuretic peptide receptor coupled guanylate cyclase (ANP-RGC) in a non-hormonal, Ca^2+^-dependent fashion and independently of ANP signaling ^29^. In cardiac and muscle cells, cGMP is generated both from nitric oxide (NO) activated soluble guanylyl cyclase and by activation of particulate guanylyl cyclases, such as ANP-RGC. Dystrophin loss results in the disruption of neuronal NO synthase localization, and muscle NO-cGMP signaling has been identified as a potential therapeutic target. A possible role for NCALD as a modifier of disease progression may be in regulating an alternate cGMP pathway and impacting the downstream signaling cascades disrupted by the loss of NO-activated cGMP synthesis. An additional target of regulatory variants in the rs3899035-tagged region may also be the zinc finger transcription factor Krüppel-like factor 10 (*KLF10*), which is functionally linked to credible SNP region by both eQTL and pCHi-C interactions. Expression of KLF10 (alias for TGF-beta inducible early gene-1) is rapidly induced by TGF-β signaling and acts as a transcriptional regulator of TGF-β responsive genes. *Klf10* knockout mice show muscle hypertrophy and fiber type distributions suggesting that KLF10 expression may be associated with muscle disease ^30^. *Klf10*^*™/™*^, *mdx* animals show dysregulated SMAD gene expression and increased fibrosis in skeletal and diaphragm muscle ^24^, suggesting a role for *KLF10* modifying effects downstream of TGF-β – LTBP4 signaling.

### Chr18 rs2061566 to PARD6G connection

#### Credible SNP set, eQTLs, and chromatin features

The chr18 SNP rs2061566 A/G variant near the chr18q subtelomere region (Figure 3A) is not in strong LD (*r*^2^ < 0.3) with other SNPs in this study or in the 1000Genomes European data set. The SNP resides within the cCRE enhancer EH38E1929501 that has prominent DHS peaks in stromal A and B components (Figure 3B) enriched for ENCODE biosamples derived from connective tissues and cells, and in the myeloid/erythroid component (Figure 3C, Table S4), which is enriched for biosamples derived from hematopoietic stem cells. Multiple, cell-type specific transcription factor (TF) footprints are evident within the DHS peak for E1929501 from both psoas muscle and CD34+ hematopoietic stem cells (Figure 3C). The top pCHi-C interaction observed in psoas muscle (Figure 3A, Table S3) connected the E1929501 enhancer to the par-6 family cell polarity regulator gamma (Partitioning Defective 6, *PARD6G*) gene. PARD6G is a member of the PAR polarity complex (PAR3-PAR6-aPKC) that participates in the control of asymmetric cell division and self-renewal of muscle satellite cells ^31^. Although *PARD6G* is not the nearest protein-coding gene to the chr18 rs2061566 SNP, we queried the STRING interaction database and observed a network (Figure 4A) connecting the PARD6/3 multiprotein complex to dystrophin through joint interactions with Mark2 (also known as PAR-1b), which is known to bind dystrophin in muscle satellite cells.

**Figure 3.**
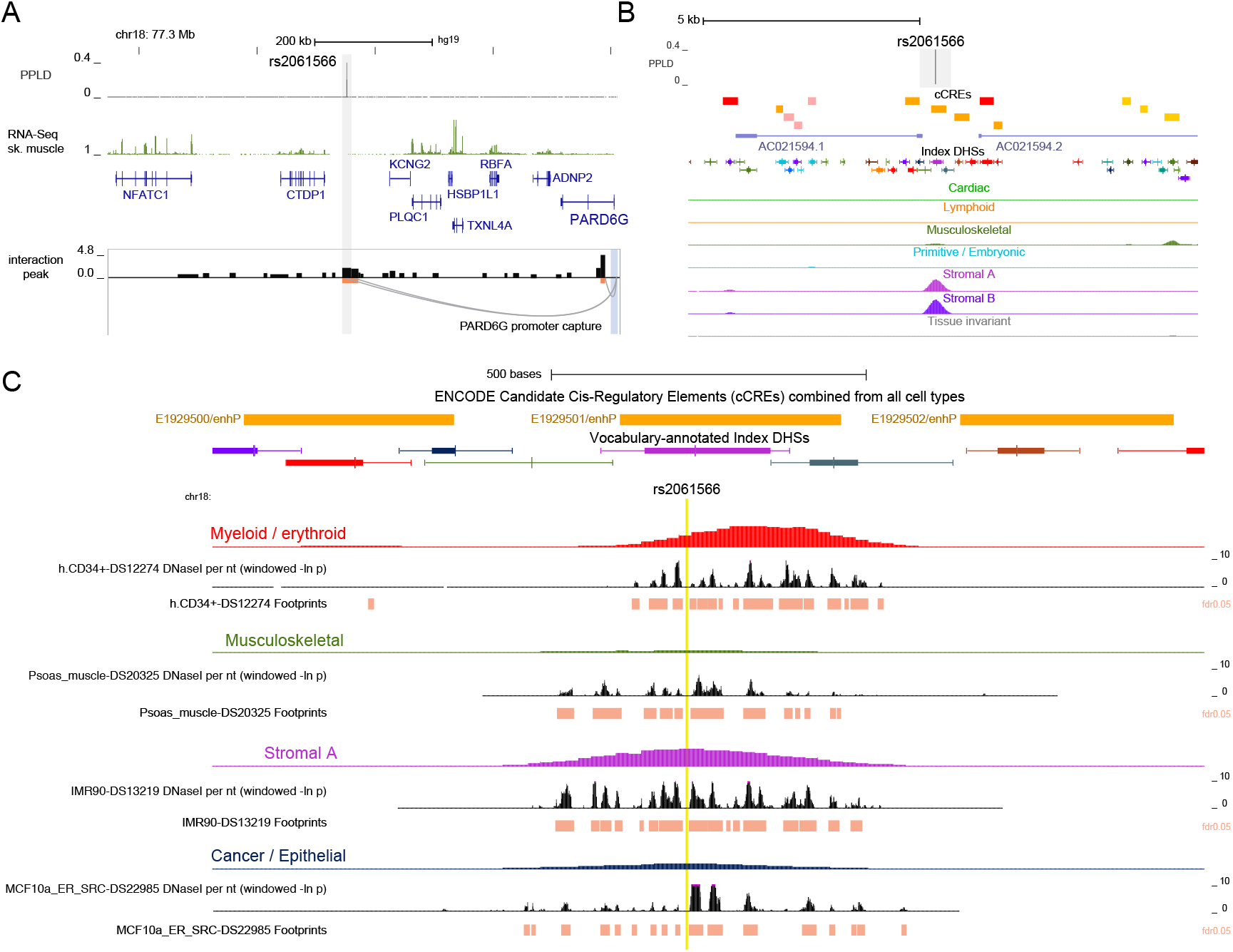
Chromatin features and TF footprints in the chr18 rs2061566 region. (A) Annotation tracks from chr18:77,150,001-78,020,000 (hg19) with the PPLD value and skeletal muscle total RNA-Seq read depths, RefSeq Select transcripts, and highlighted credible SNP region similar to Figure 1. The promoter capture Hi-C chromatin interactions from psoas muscle using the *PARD6G* chr18:77,995,738-78,007,496 promoter fragment. (B) The credible SNP region (grey bar) from (A) with annotation tracks for ENCODE cCREs, vocabulary-annotated index DHSs elements, and biologically labeled component meta-DNase I tracks from the top 15 ENCODE biosamples most strongly associated with each component. C) Annotation tracks for ENCODE candidate cis-Regulatory Elements (cCREs), vocabulary-annotated index DHSs elements, and biologically labeled component DNase I tracks as in (B). DNase I cleavage pattern from Vierstra et al. (2020), showing DNaseI per-nucleotide cleavage and the digital TF footprints within the DHS at an FDR cutoff = 0.05. The location of rs2061566 is highlighted by the light-yellow bar.

**Figure 4.**
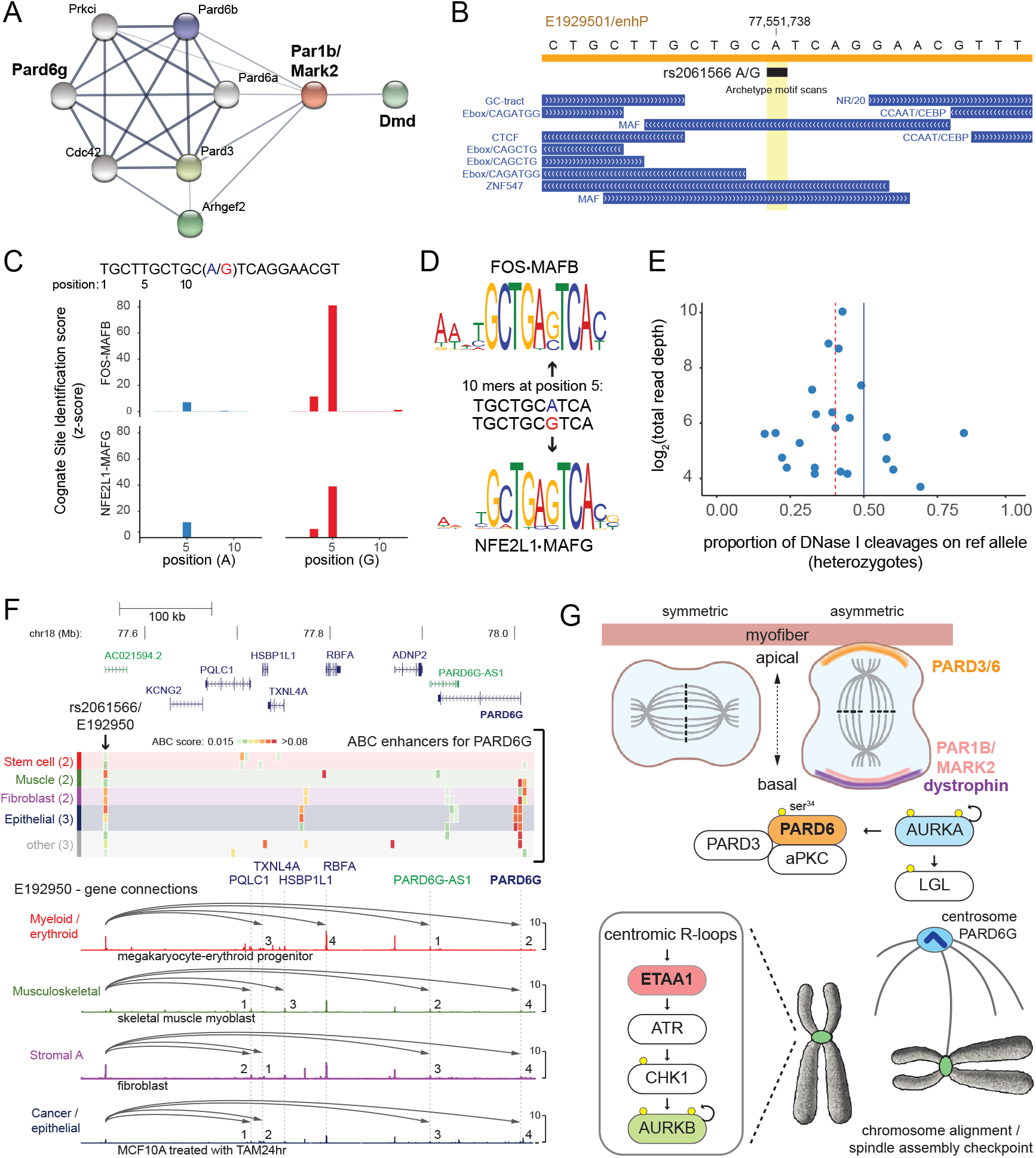
The rs2061566 SNP alters an enhancer linked to *PARD6G*. (A) Two-layer protein network from the STRING database using *Mus musculus* Mark2 as the query. Nodes are proteins and edges are the connection strengths using ‘Experiments’, ‘Databases’ and ‘Textmining’ as interaction data sources. (B) Sequence features in the rs2061566 region, including the cCRE E192950 enhancer (orange) and transcription factor binding motifs (blue) assigned from index DHS TF footprints. (C) Cognate site identification scores predict increased binding of FOS-MAFB and NFE2L1-MAFG heterodimers to the rs2061566 A allele (blue) versus the G allele (red). (D) Representative MEME motifs are shown for the predicted bZIP TF heterodimers. (E) Scatterplot of rs2061566 allelic imbalance from heterozygous samples indicate increased TF footprinting on the alternate allele. (F) RefSeq transcript annotation tracks from chr18: 77,500,001-78,020,000 (hg19) are shown with protein coding transcripts in blue and lincRNAs in green. The heatmap shows enhancers predicted to regulate *PARD6G* (ABC score >= 0.015, heatmap color scale for ABC score bins), with the location of rs2061555/E1929501 enhancer indicated with an arrow. The top 4 ABC E1929501 enhancer-gene predictions are indicated by arcs from the E1929501 enhancer to the contacted promoter over tracks for the biologically labeled DHS component meta-DNase I tracks. These arcs are numbered by their ABC score rank (Table S8) in megakaryocyte-erythroid progenitor cells (Myeloid/erythroid DHS component), in skeletal muscle myoblasts (Musculoskeletal DHS component), in IMR90 fibroblasts cells (Stromal A DHS component), and in MCF 10A cells transformed with v-Src (Cancer/epithelial DHS component). (G) Model for postulated roles ETAA1 and PARD6G in control of symmetric/asymmetric cell division in satellite cell niche. Arrows indicate signaling cascades with phosphorylation events indicated by yellow circles.

#### Functional evidence linking rs2061566 to E1929501 enhancer activity

We used ENCODE transcription factor (TF) footprints ^32^ to assess whether rs2061566 disrupted a regulatory protein binding site within the E1929501 enhancer. Figure 4B shows that rs2061566 intersects overlapping small MAF (musculoaponeurotic fibrosarcoma) binding sites for homo- and heterodimeric basic region leucine zipper-type (bZIP) TFs. The rs2061566 allelic effect on bZIP TF binding was tested using empirical DNA binding z-scores (CSI scores = cognate site identification 10mer intensity scores) from 109 bZIP TF homo- and heterodimeric pairs ^33^. We ranked CSI scores using a sliding window across a 21 bp. sequence centered on rs2061566 and observed a maximum CSI binding score for FOS-MAFB bZIP heterodimers at a position containing the alternate rs2061566 G allele (Figure 4C). Similar binding specificity was also observed for NFE2L1-MAFG heterodimers and the binding specificities of both bZIP TF heterodimers at position 5 were consistent with their representative heterodimer motifs (Figure 4D). We tested this predicted gain-of-function effect by examining digital TF footprinting data from ENCODE tissue/cell biosamples heterozygous for rs2061566 (Table S5). The allelic imbalance observed for reads overlapping the rs2061566 site (Figure 4E) suggests that the rs2061566 G allele is associated with increased bZIP TF binding *in vivo*.

The cell type specificity and strength of the E1929501 enhancer – *PARD6G* promoter interaction relative to other promoters in the region was examined using the activity-by-contact (ABC) model of enhancer-promoter maps from 131 human cell types and tissues ^34^. The ABC score measures enhancer–gene connections based on enhancer activity derived from DHS and H3K27ac chromatin data and weighted by the frequency of normalized Hi-C chromatin interactions with individual promoters. Using a minimum threshold ABC score >= 0.015, the E1929501 enhancer contacted 11 gene promoters in 18 (out of 131) samples (Table S6), and the E1929501 – *PARD6G* promoter connection was observed in 12 of these 18 samples. A total of 84 ABC enhancer – *PARD6G* promoter connections were observed in these 131 samples (Figure 4F, Table S7), and we examined ABC enhancer scores from cell types that are representative for the components with the most prominent E1929501 DHS peaks, including megakaryocyte-erythroid progenitor stem cells (myeloid/erythroid), skeletal muscle myoblasts (musculoskeletal), IMR90 fibroblasts (stromal A), and transformed MCF10A epithelial cells (cancer/epithelial). The top four E1929501 enhancer-gene ABC scores in each of these cell types included the *PARD6G* promoter, with the top-ranked *PARD6G* score observed in myeloid/erythroid progenitor stem cells (Figure 4F, Table S8).

#### *PARD6G* as a plausible modifier gene

The location of rs2061555 in a prominent cCRE enhancer that has strong activity-by-contact chromatin links to the *PARD6G* promoter in stem cells and the altered transcription factor binding to the heterodimeric MAF motif was the most direct evidence we observed for a functional SNP effect. Single cell transcriptome analysis of muscle stem cells from adult mouse hindlimb identified the bZIP transcription factor MAFF as one of the top-75 differentially expressed genes in the muscle stem/progenitor and myonuclei clusters ^35^. *Maff* expression is highest in quiescent MuSCs suggesting a potential functional effect through rs2061566-altered binding of MAFF heterodimers to this cCRE distal enhancer in these cell types. Stem cell (including satellite cell) polarity is driven by the localization of this complex (PAR3-PAR6-aPKC) to the apical pole, along with the localization of the PAR1 complex to the basilar pole ^36^. These complexes are critical for the localization of centrosomes central to the asymmetric cell division resulting in a population of self-renewed satellite cells and a population of *Myf-*expressing cells leading to myofiber regeneration ^36^. The *DMD* gene is expressed in satellite cells ^31^ and dystrophin interacts with the Mark2 (or PAR-1b) protein via specific interaction within the 8^th^ and 9^th^ spectrin-like repeats within its central rod domain ^37^. Although muscle from DMD patients shows increased numbers of satellite cells, particularly in association with type 1 fibers in biopsies from patients with increased age ^38^, polarity and associated asymmetric division into regenerative cells is impaired in DMD muscle^36,39^.

## Discussion

Analysis of 6 loci with PPLD scores of ≥0.4 identified multiple genes suitable for further exploration, based upon multiple lines of evidence for plausibility for a biologic role for the identified SNPs in regulating genes or pathways potentially relevant to muscle biology. For each region, we systematically applied a search for features indicative of functional regulatory variation, for evidence of associated gene regulation by eQTL analysis, and for evidence of direct physical interaction via Hi-C interactions. This uniform approach provided multiple concordant lines of evidence supporting each of the candidate genes, each of which may play a role in muscle function or pathology. Although the PPLD approach suggested a distinct set of modifier loci from previously described candidates, the overlap of gene functions in these data sets suggests that recurrent biological pathways are affected.

Overlapping functions in muscle stem cell maintenance and division for *ETAA1* and *PARD6G* suggests a model connecting these two genes to dystrophin loss in satellite cells (Figure 4G). Impairments of the MuSC population in dystrophic muscle have been attributed to exhaustion due to constant activation, to defects in asymmetric cell division and reduction in myogenic precursor cells caused by an intrinsic lack of dystrophin in MuSCs, and to differences in the stem cell niche ^31^. In animal models of DMD, such as the Golden Retriever Muscular Dystrophy dog, increased expression of the Notch ligand Jagged1 is associated with a mild phenotype that is postulated to result from increased proliferation of myogenic precursors from MuSCs ^7^. In an *mdx* mouse model engineered with defective telomerase activity, telomere shortening was associated with increased replicative senescence of *mdx* MuSCs and increased disease severity ^40^. In that model, telomere shortening induced markers of the DNA damage response which impaired MuSC differentiation into myotubes *in vitro* by interfering with MYOD-mediated activation of myogenic gene expression ^41^. The SNP effects on *ETAA1* expression may suggest a vulnerability in rapidly proliferating MuSCs that activates the ETAA1-ATR signaling pathway during S and M cell cycle progression due to elevated replicative stress or incompletely replicated loci.

Recently, it has been shown that signaling through the Aurora A kinase (AURKA) cascade can rescue some of the polarity and asymmetric satellite cell division defects observed in *mdx* mice ^42^. Stimulation of mouse Aurka activity through the EGFR signaling pathway increases the proportion of asymmetric cell divisions, while pharmacological inhibition of Aurka shifts the balance towards symmetric stem cell division. Prior work established that Aurora A kinase phosphorylates the ser^34^ residue of PARD6G and that this is a key step in regulating the composition of the PAR complex including the activation of αPKC and the release of the Lgl subunits. Independent of its role in the PAR complex, PARD6G also has a unique role in the two centrioles of the centrosome, where PARD6G preferentially associates with the older mother centriole to regulate centrosomal protein composition, including the formation of spindle poles that control the accurate segregation of DNA into the two daughter cells ^43^. Thus, the regulatory SNPs identified for *ETAA1* and *PARD6G* may play multiple roles in cell polarity and asymmetric division which impact the response to dystrophin loss in satellite cells. The degree of impairment to the eventual deterioration of muscle integrity (and ultimately function) is arguably understudied and may be of increased importance to understand as microdystrophin constructs in gene therapy trials do not encode the Mark2-binding spectrin repeats. Along with the fact that muscle stem cells are poorly transduced *in vivo* by current AAV vectors and would poorly express transgene with the muscle-specific promoters currently in use, this raises the possibility that microdystrophin gene therapy will not correct fundamental muscle stem cell defects important to muscle maintenance.

## Summary

Our results suggest new plausible candidates for genetic modifiers of DMD disease severity as measured by LOA. Studies that examine the interaction between large-effect monogenic mutations and small-effect modifier variants, such as the one described here, are few in number. It is notable that the associations of common SNPs detected with this study were all non-coding variants, consistent with the majority of GWAS-associated variants linked to human traits. Recent studies have observed that the polygenic background as measured by risk scores for common diseases such as coronary heart disease, breast cancer, and colon cancer, modifies the disease penetrance of hereditary forms of these diseases ^44^, supporting a ‘many modifiers’ model for monogenic disease phenotypes. That recurrent biological pathways are evident between the previously described DMD modifiers and the new set of potential modifiers we report here suggests that therapeutic strategies based on these pathways requires further understanding of their common physiological action in the context of the disease-causing mutation.

## Data Availability

All data produced in the present study are available upon reasonable request to the authors

## ETHICAL APPROVAL

The research involving human subjects has been performed in accordance with the Declaration of Helsinki and was approved by the Nationwide Children’s Hospital Institutional Review Board (IRB) under approval 0502HSE046. Informed consent was obtained from all subjects.

## ACKNOWLEDGMENTS

This work was supported by the National Institutes of Health (NINDS NS085238) to KMF, RBW, and VJV. The Genotype-Tissue Expression (GTEx) Project was supported by the Common Fund of the Office of the Director of the National Institutes of Health, and by NCI, NHGRI, NHLBI, NIDA, NIMH, and NINDS. The data used for the analyses described in this manuscript were obtained from the GTEx Portal using the GTEx Analysis Release V8 (dbGaP Accession phs000424.v8.p2). The authors wish to acknowledge the many members of the United Dystrophinopathy Project consortium who participated in the enrollment of subjects in the original historical UDP database.

## AUTHOR CONTRIBUTIONS

R.B.W., V.J.V. and K.M.F conceived and designed the study. R.B.W., V.J.V., D.M.D., M.W., R.A., L.A., M.M-C., K.M.F., J.B., SC.S., and the UDP Investigators acquired and/or analyzed the data. R.B.W., V.J.V., P.T.M., and K.M.F. drafted the manuscript, which was reviewed and revised by all of the named authors.

## Supporting information

### Supplemental Methods

**Table S1**. SNP annotation of PPLD credible sets

**Table S2:** SNP-gene pairs for cis-eQTLs from data sources aggregated by the FUMA SNP2GENE process.

**Table S3**. Promoter capture Hi-C chromatin interactions 1 within PPLD credible set ranges.

**Table S4**. Candidate SNP overlap with ENCODE DNaseI Hypersensitive Sites (DHS) Regulatory Vocabulary1 and Candidate Cis-Regulatory Elements (cCREs).

**Table S5**. Allelic imbalance from DNase I cleavage data of ENCODE biosamples heterozygous at rs2061566.

**Table S6**. Activity by contact EH38E192950 enhancer-gene mapping in 18 cell types and tissues.

**Table S7**. Activity by contact enhancer-PARD6G gene mapping in 12 cell types and tissues.

**Table S8**. Activity by contact rs2061566 (chr18:77551738) EH38E1929501 enhancer-gene mapping.

